# SPLASH: A Benchtop Platform for Accessible, Ultrasensitive Quantification of Plasma Biomarkers in Alzheimer’s Disease

**DOI:** 10.64898/2026.02.21.26346786

**Authors:** Noah Elder, Hien Nguyen, Jason Wan, Taylor P. Johnson, Marissa Lee, Christopher Ng, Jennifer S. Yokoyama, Robert Lin

**Affiliations:** Taudia, Inc., San Carlos, United States; Edward and Pearl Fein Memory and Aging Center, Department of Neurology, Weill Institute for Neurosciences, University of California, San Francisco, CA, USA; Department of Radiology and Biomedical Imaging, University of California, San Francisco, CA, USA

**Author notes:** Corresponding author –. These authors contributed equally to this work (Noah Elder, Hien Nguyen).

## Abstract

Blood-based biomarkers have emerged as a promising tool for the detection and monitoring of neurodegenerative diseases such as Alzheimer’s disease (AD), yet broad implementation of ultrasensitive protein quantification remains constrained by reliance on specialized instrumentation and centralized laboratory infrastructure. Here we present SPLASH (Solid Phase Ligation Assay with Single wasH), an ultrasensitive proximity ligation assay platform that achieves sub-pg/mL sensitivity using only standard benchtop qPCR equipment. We developed five assays targeting Alzheimer’s disease biomarkers – pTau-217, Aβ1–40, Aβ1–42, neurofilament light chain (NfL), and glial fibrillary acidic protein (GFAP) – with limits of detection ranging from 0.0005 to 0.119 pg/mL. Direct comparison with Simoa demonstrated high concordance (R^2^ = 0.95) for plasma pTau-217 quantification across AD-positive and AD-negative samples. We further established compatibility with dried plasma spot samples, enabling decentralized collection and quantitation without cold-chain storage. A multiplexed five-analyte panel was applied to 69 plasma samples, revealing heterogeneous biomarker profiles consistent with AD-associated patterns. By eliminating dependencies on proprietary instrumentation, SPLASH facilitates broad implementation of ultrasensitive protein quantification for neurodegenerative disease research and diagnostics.

## Introduction

Blood-based biomarkers have the potential to revolutionize neurological disease management by enabling early detection and longitudinal monitoring [1–3]. Compared with other methods, such as cerebrospinal fluid (CSF) collection or brain positron emission tomography (PET) imaging, blood sampling is less invasive, scalable and largely accessible [2]. Blood-based biomarkers have already been shown to correlate with key pathological processes in neurological disorders like Alzheimer’s Disease (AD) [4–11], Parkinson’s Disease (PD) [3,12,13], and traumatic brain injury (TBI) [14,15]. Recent FDA approval of blood-based tests for AD underscores their clinical relevance, helping enable biologically-based diagnostics [16–20]. However, broad implementation of blood-based biomarker testing remains limited by the platform-level constraints of ultrasensitive protein quantitation.

While identification of relevant blood-based biomarkers has been accelerated by proteomic discovery platforms such as mass spectrometry, Olink’s Proximity Extension Assay (PEA) [21], and Alamar Biosciences’ Nucleic Acid Linked Immuno-Sandwich Assay (NULISA) [22], translation into clinical validation studies often narrows to a limited number of biomarkers measured on low-plex platforms such as Quanterix’s Simoa [23], Roche’s Elecsys [24], or Fujirebio’s Lumipulse [16]. However, these methods require substantial capital investment and specialized instrumentation. Implementation analyses and global health assessments emphasize that successful deployment further depends on laboratory infrastructure, trained personnel, and harmonized pre-analytical workflows that remain unevenly distributed across health systems [25,26], contributing to persistent diagnostic access gaps globally, particularly in low- and middle-income countries (LMICs) [27] (Figure 1a). Current ultrasensitive biomarker platforms require centralized laboratories and specialized instrumentation (Fig. 1b, top), potentially limiting deployment in resource-limited settings.

**Figure 1.**
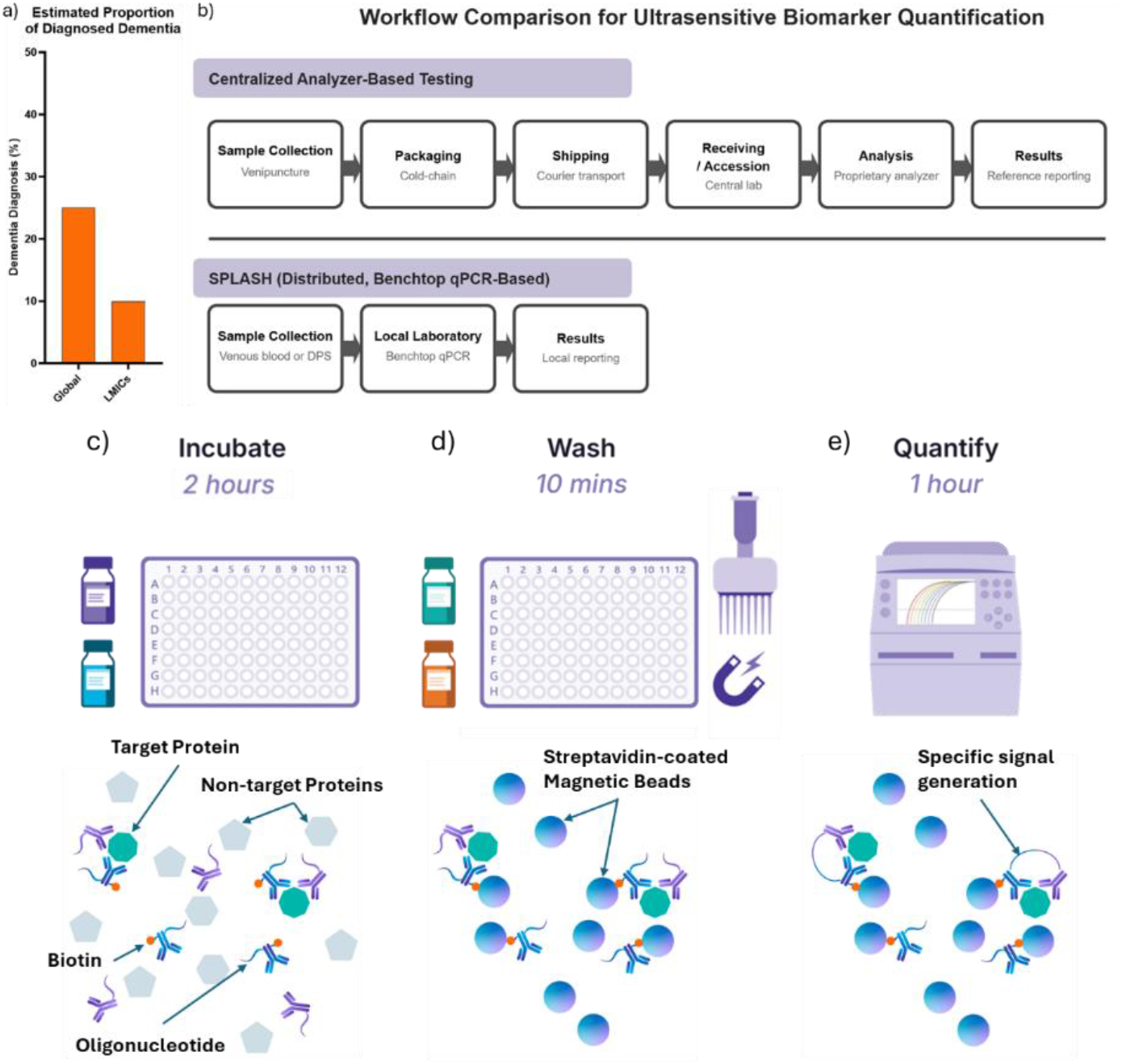
SPLASH (Solid Phase Ligation Assay with Single wasH) enables decentralized ultrasensitive biomarker quantification. **(a)** Estimated proportion of dementia cases receiving formal diagnosis globally (∼25%) versus LMICs (∼10%). **(b)** Workflow comparison for ultrasensitive biomarker quantification. Top: Centralized analyzer-based testing requires cold-chain transport to specialized laboratories. Bottom: SPLASH enables local testing using venous blood or dried plasma spots (DPS) with standard qPCR instrumentation. **(c-e)** SPLASH workflow: **(c)** Antibody probes conjugated to oligonucleotides bind target protein during incubation. **(d)** Streptavidin-coated magnetic beads capture immunocomplexes followed by magnetic wash. **(e)** qPCR amplification and quantification of ligated oligonucleotides.

To overcome these limitations, we developed SPLASH (Solid Phase Ligation Assay with Single wasH), an immunoassay platform that enables ultrasensitive protein quantitation without specialized instrumentation (Fig. 1b, bottom). SPLASH is a wash-based proximity-ligation workflow consisting of three steps: (1) homogeneous incubation to form immunocomplexes (Figure 1b), (2) a magnetic bead wash to remove unbound material (Figure 1c), and (3) qPCR-based quantification (Figure 1d). The assay uses two antibody probes conjugated to oligonucleotides; one of the antibodies is also conjugated to biotin. The antibody probes bind to the target protein during incubation. After immunocomplex formation, streptavidin-coated magnetic beads are added to capture the complexes, via biotin-streptavidin interaction. Using a magnetic rack, the beads are held in the wells during washing to remove excess probes and sample matrix. The oligonucleotides in the captured immunocomplexes are then ligated and quantified by qPCR, with absolute concentrations calculated from a standard curve.

This design achieves the sub–pg/mL sensitivity required for low-abundance biomarkers such as plasma phosphorylated tau-217 (pTau-217), while reporting absolute concentrations in conventional units (i.e. pg/mL), facilitating cross-platform and cross-study comparability. Because its simplified workflow removes dependencies on expensive proprietary instruments, SPLASH significantly lowers both the capital investment and technical expertise required, thus facilitating deployment of ultrasensitive protein quantitation beyond centralized laboratories.

In this study, we first benchmarked SPLASH performance against the Simoa HD-X platform, demonstrating strong concordance (R^2^=0.95) for plasma pTau-217 quantitation on 20 plasma samples. We then established feasibility of deployment in resource-limited settings by successfully quantifying pTau-217 and Aβ1–42 levels in Dried Plasma Spots (DPS). Finaly, we extended SPLASH into a multiplexed, 5-target panel to detect progressive biomarker profile changes in 69 plasma samples. Taken together, these results demonstrate that SPLASH supports scalable implementation of accurate blood-based biomarker quantitation for neurological disease across diverse settings.

## Results

### SPLASH demonstrates strong analytic performance

To evaluate the analytical performance of SPLASH, we developed five assays targeting Alzheimer’s disease-associated biomarkers: pTau-217, amyloid β1–40 (Aβ1–40), amyloid β1–42 (Aβ1–42), neurofilament light chain (NfL), and glial fibrillary acidic protein (GFAP). We assessed assay performance by determining the lower limit of detection (LLOD), lower limit of quantification (LLOQ), inter- and intra-plate repeatability, plasma detectability, and correlation with a commercially available ultrasensitive platform (Simoa, Quanterix).

All five assays demonstrated sub-pg/mL sensitivity with LLOD values ranging from 0.0005 to 0.119 pg/mL (Aβ1–40: 0.119 pg/mL, Aβ1–42: 0.0005 pg/mL, pTau-217: 0.001 pg/mL, NfL: 0.077 pg/mL, GFAP: 0.075 pg/mL) and LLOQ values from 0.007 to 2.5 pg/mL (Aβ1–40: 1.03 pg/mL, Aβ1–42: 0.283 pg/mL, pTau-217: 0.007 pg/mL, NfL: 2.19 pg/mL, GFAP: 2.5 pg/mL) (Figure 2a). Overall mean intra-plate coefficient of variation (CV) was 4.76% across all targets and 2 different plasma samples (2.29% to 8.57%) (Figure 2b).

**Figure 2.**
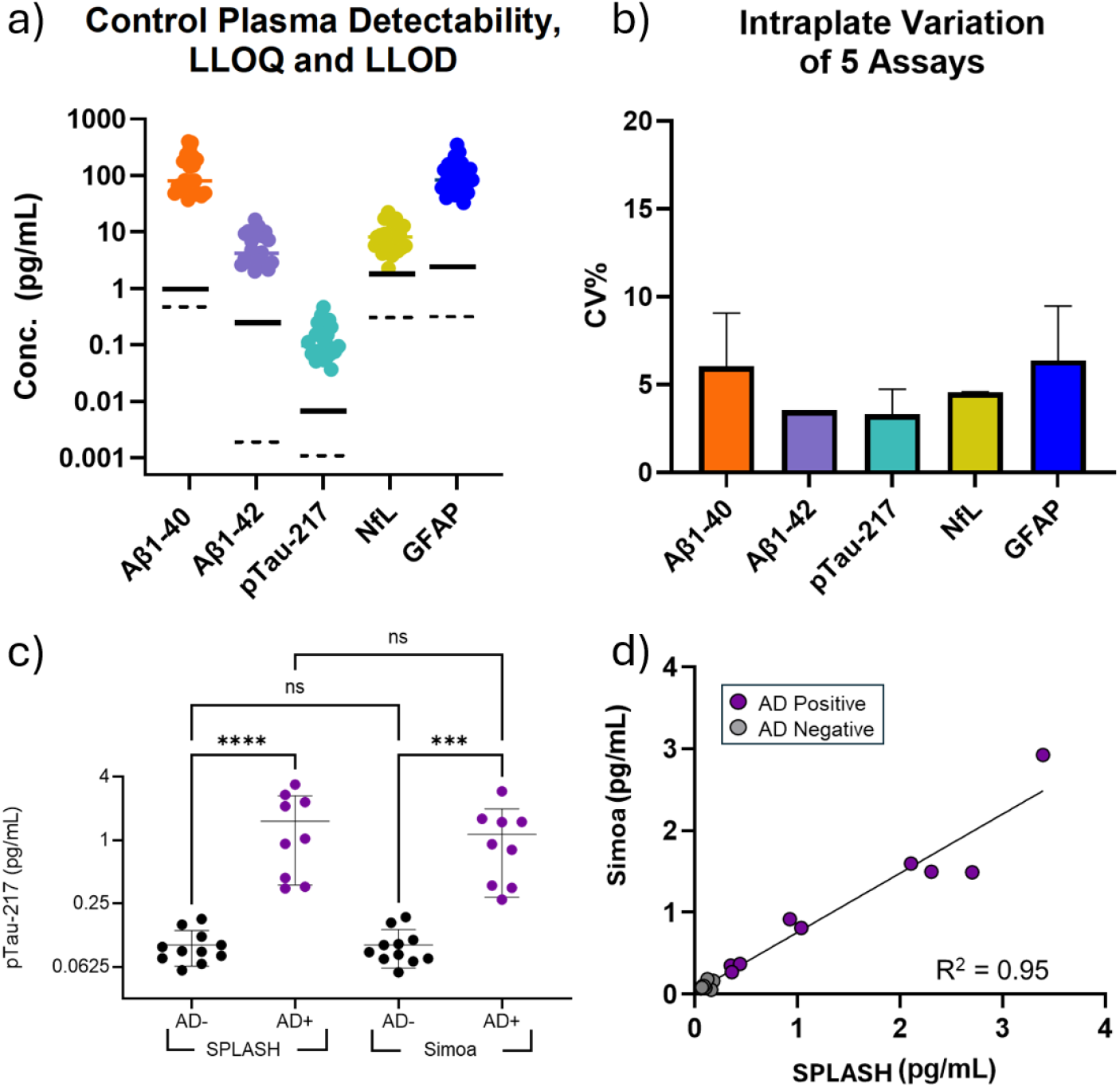
Analytical performance and performance confirmation of the SPLASH pTau-217 assay. **(a)** Control plasma detectability for five biomarkers (Aβ1-40, Aβ1-42, pTau-217, NfL, GFAP) relative to lower limit of quantification (LLOQ, solid line) and detection (LLOD, dashed line). **(b)** Intra-plate reproducibility represented as CV%. Vertical bars are standard deviations. **(c)** pTau-217 concentrations in AD-positive and AD-negative samples measured by SPLASH and Simoa (****p<0.0001, ***p<0.001, ns=not significant). **(d)** Correlation between SPLASH and Simoa measurements (R^2^=0.95).

All 24 control plasma samples yielded concentrations above the LLOQ. Across all assays, median and mean concentrations were at least 80-fold above the LLOD, with pTau-217 representing the lowest margin (Figure 2a). Median and mean concentrations exceeded the LLOQ by at least 10-fold for all assays except NfL, which showed a median of 3.7-fold above LLOQ.

To benchmark SPLASH against an established platform, we quantified pTau-217 in 20 commercially sourced plasma samples (11 AD-negative, 9 AD-positive) using both SPLASH and Simoa. Both assays clearly distinguished AD-positive from AD-negative samples using a single threshold (SPLASH: p<0.001, Simoa: p<0.001), with no significant difference in measurements between platforms within each diagnostic group (Figure 2c). Plasma pTau-217 levels measured by the two platforms were highly correlated (R^2^ = 0.95, Figure 2d).

### Dried plasma spots to enable global sample collection and quantitation

To assess compatibility with decentralized sample collection and handling conditions, we evaluated SPLASH performance using dried plasma spot (DPS) samples. Throughout collection, processing, and analysis, samples were handled without cold-chain storage to reflect decentralized handling conditions. Each sample was collected onto DPS cards that contain two collection discs each. For all 16 samples, each disc was tested in duplicate using a duplex pTau-217/Aβ1–42 assay with all samples demonstrating signal above background for both pTau-217 and Aβ1–42 (Figure 3a). Coefficients of variation (CV) across replicate wells from individual SEP10 DPS discs were low (pTau-217 mean CV = 6.0%, Aβ1–42 mean CV = 6.3%), indicating high technical precision. In contrast, values across multiple DPS discs per sample exhibited higher variability (pTau217 mean CV = 34.7%, Aβ1–42 mean CV = 17.8%), suggesting high degree of variability in DPS extraction efficiency (Table 1).

**Figure 3.**
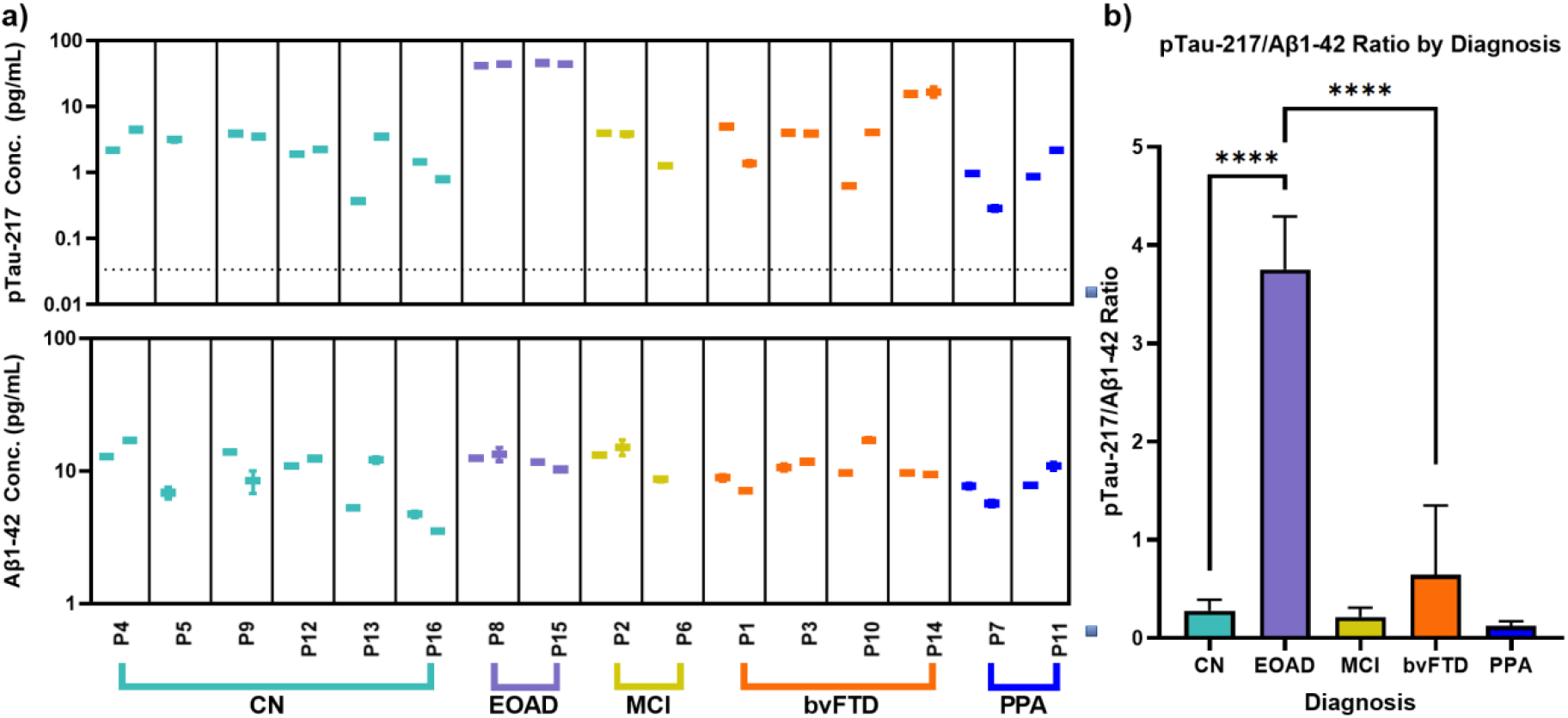
pTau-217 and Aβ1–42 plasma concentrations and pTau-217/Aβ1–42 ratio across diagnostic groups. **(a)** Individual pTau-217 (top) and Aβ1-42 (bottom) concentrations for cognitively normal (CN), early-onset Alzheimer’s disease (EOAD), mild cognitive impairment (MCI), behavioral variant frontotemporal dementia (bvFTD), and primary progressive aphasia (PPA) samples. Dotted line indicates pTau-217 background. Aβ1–42 background below 1pg/mL. **(b)** pTau-217/Aβ1-42 ratio by diagnostic group showing significantly elevated ratios in EOAD samples (****p<0.0001).

Calculated pTau217/Aβ1–42 ratios in the early-onset Alzheimer’s disease (EOAD) group were significantly higher than all other diagnostic groups (p < 0.0001), consistent with established disease-associated biomarker patterns [28,29] (Figure 3b). Mean cohort ratios were 0.28 ± 0.11 (clinically normal, CN), 3.75 ± 0.55 (EOAD), 0.212 ± 0.091 (mild cognitive impairment, MCI), 0.645 ± 0.706 (behavioral variant frontotemporal dementia, bvFTD), and 0.12 ± 0.048 (primary progressive aphasia, PPA). No statistically significant differences were observed between CN, MCI, bvFTD, or PPA groups. The calculated pTau-217/Aβ1–42 ratio showed within-disc CV of 8.1% and between-disc CV of 29.6% (Table 1). Together, these results demonstrate that DPS-based sample handling can be combined with the SPLASH workflow to enable sensitive quantitation of Alzheimer’s disease biomarkers under decentralized handling conditions.

**Table 1.** Mean Coefficient of Variation (CV) of Measured Biomarker Levels for All Samples.

|  | Within-disc CV (%) | Between-disc CV (%) |
| --- | --- | --- |
| **pTau-217** | 6.0% | 34.7% |
| **Aβ1–42** | 6.3% | 17.8% |
| **pTau-217/Aβ1–42** | 8.1% | 29.6% |

Within-disc: technical replicates from the same DPS disc; Between-disc: all replicates from different DPS discs.

### Multiplexed biomarker profiling reveals progressive changes

We next evaluated SPLASH performance in a multiplexed assay format using a panel of AD-associated biomarkers, as a multi-analyte approach provides more comprehensive information for each sample. We created a 5-biomarker panel consisting of two duplex assay, pTau-217 with Aβ1–42 and NfL with GFAP with single-plex Aβ1–40 and tested 69 commercially sourced samples. Of these, 50 were uncharacterized plasma samples from individuals aged 50–80 years and 19 were AD-positive plasma samples. All five analytes were successfully detected across all samples, with median concentrations of 0.42 pg/mL for pTau-217, 2.46 pg/mL for Aβ1–42, 5.81 pg/mL for Aβ1–40, 0.48 pg/mL for NfL and 14.7 pg/mL for GFAP (Table 2). All median signals were at least 40X above their respective background signals. While the measured level of pTau-217, NfL and GFAP were used as is, the ratio of Aβ1–42-to-Aβ1–40 (Aβ1–42/Aβ1–40) was used in subsequent biomarker categorization [30,31].

A two-cutoff system was used to categorize biomarker levels into normal, intermediate and abnormal categories. Cutoffs were determined using an AD-anchored approach based on the biomarker distribution of confirmed AD samples. This was done as the uncharacterized samples cannot be assumed to be healthy controls. Details regarding the cutoffs are described in the Methods section. In the AD-positive samples, changes in the Aβ1–42/Aβ1–40 ratio were consistently observed, followed by elevations in GFAP and pTau-217, with increases in NfL generally appearing later (Figure 4a). This pattern is consistent with established biomarker progression models and aligns with recent plasma proteomic data demonstrating associations among these core AD biomarkers [32,33] (Figure 4a).

**Figure 4.**
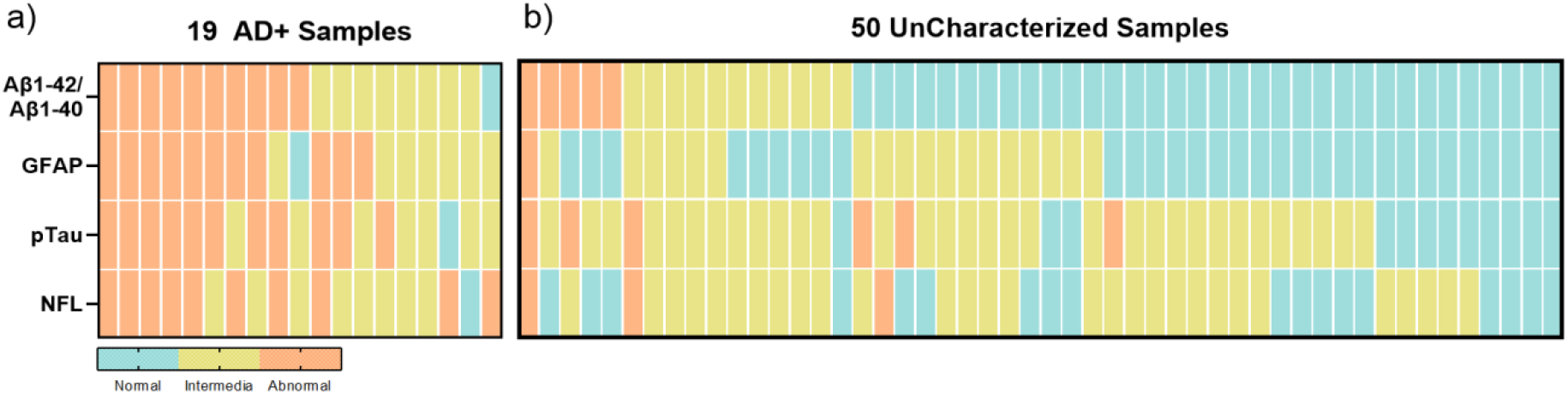
Multiplex quantification and profiling of Alzheimer’s disease biomarkers by SPLASH. **(a)** Heatmap of Aβ1– 42/Aβ1–40 ratio, GFAP, pTau-217, and NfL levels in AD-positive samples (n = 19). Biomarker levels were categorized as normal (blue), intermediate (yellow), or abnormal (orange) using AD-anchored cutoffs as described in Methods. **(b)** Biomarker profiles in uncharacterized plasma samples from individuals aged 50–80 years (n = 50).

Notably, a number of uncharacterized plasmas also presented abnormal biomarker levels (38-72% depending on biomarker) (Figure 4b). This is consistent with the age range of the cohort (50-80 years) and published estimates of preclinical AD prevalence in this age group (20-40%) [34,35].

**Table 2.** Detectability of 5 biomarkers across plasma samples.

|  | Mean<br>(pg/mL) | Median<br>(pg/mL) | Median Signal Folds<br>Above Bkgd | Detection |
| --- | --- | --- | --- | --- |
| pTau-217 | 0.48 | 0.42 | 59 | 100% |
| Aβ1–42 | 4.15 | 2.46 | 74 | 100% |
| Aβ1–40 | 11.9 | 5.81 | 277 | 100% |
| NfL | 0.56 | 0.48 | 58 | 100% |
| GFAP | 17.9 | 14.7 | 149 | 100% |

## Discussion

Our analytical testing results demonstrate that SPLASH achieves sub-pg/mL sensitivity across all five neurological biomarkers, with LLOD values comparable to or exceeding those of established ultrasensitive platforms. Notably, even pTau-217, which showed the narrowest margin at 88-fold above LLOD, remained well within the quantifiable range. Margins above LLOQ were similarly robust, exceeding 10-fold for all assays except NfL (3.7-fold). The single-digit mean CV for intra-plate measurements (4.76%) suggests excellent reproducibility, particularly for manual workflow with no automated liquid handling or plate processing. This satisfies the level of precision required to facilitate longitudinal studies monitoring disease progression and for multi-site studies where plate-to-plate consistency is essential.

In a direct platform comparison SPLASH demonstrated comparable ability to discriminate AD-positive from AD-negative samples as Simoa, with both platforms achieving clear separation using a single threshold and showing no significant difference in measurements within diagnostic groups. The strong correlation between platforms (R^2^ = 0.95) is particularly notable given that the two systems utilize different antibodies, distinct chemistries, and independent reference materials for the generation of standard curves. This cross-platform concordance demonstrates that SPLASH can produce clinically meaningful results comparable to an established commercial platform widely used in AD research and clinical trials. This is consistent with the current state of the field, in which multiple technologies – including Simoa, Meso Scale Discovery (MSD), and Lumipulse, among others – demonstrate comparable pTau-217 quantitation performance despite methodological differences [36–38].

To demonstrate SPLASH’s viability in resource-limited settings, we applied the platform to dried plasma spot (DPS) samples in a proof-of-concept study. While SPLASH achieved low intra-disc CVs, indicating high technical assay precision, the elevated inter-disc variability strongly suggests that further optimization of both sample collection protocols and DPS extraction methodology is required. Notably, the pTau-217:Aβ1–42 ratio demonstrated lower between-disc variability (29.6%) compared to pTau-217 alone (34.7%) and fell below the independent error prediction (39%). This suggests that the ratiometric approach can mitigate the variation introduced by pre-analytical effects that similarly affect both analytes, consistent with observations in CSF where Aβ1–42/1–40 ratios show improved stability compared to individual measurements [39]. The reduced variability of biomarker ratios may be particularly valuable for DPS-based testing, where pre-analytical factors (sample collection, drying conditions, storage) introduce additional sources of variation [40].

To further demonstrate the scalability of the SPLASH platform, we assembled a 5-biomarker panel measuring pTau-217, Aβ1–42, Aβ1–40, NfL, and GFAP. This combination has been widely studied in the context of Alzheimer’s disease, with each biomarker reflecting distinct pathophysiological processes along the disease continuum [8,32,33]. Changes in the Aβ1–42/Aβ1–40 ratio reflect amyloid aggregation and plaque formation, the earliest detectable pathological change in AD. Elevated GFAP levels indicate astrocytic activation in response to amyloid pathology, while increased pTau-217 is linked to tau pathology and correlates strongly with amyloid burden. NfL elevation reflects axonal injury and neurodegeneration, which typically emerge later in disease progression. The observed biomarker patterns across samples were consistent with this progressive pathophysiological cascade, even in the absence of formal disease staging information.

Notably, multiple samples from the commercially sourced “uncharacterized” cohort presented biomarker profiles similar to those observed in confirmed AD-positive samples. Given the age range of these samples (50-80 years), it is plausible that some represent individuals with undiagnosed preclinical AD or other neurodegenerative conditions, consistent with the prevalence of AD pathology in this age group [34,35]. This observation highlights the importance of well-annotated cohorts in biomarker validation studies; commercially sourced plasma samples marketed as “healthy” or “normal” often lack rigorous clinical characterization and may harbor undetected pathology.

To address this limitation, we employed an AD-anchored cutoff approach, establishing thresholds based on confirmed disease samples rather than assuming the commercial cohort was disease-free. This strategy provides a conservative biomarker stratification when validated negative controls are unavailable and reflects real-world clinical scenarios where diagnostic thresholds must discriminate pathology from a heterogeneous population. In contrast, the AD-negative cohort used in the Simoa concordance analysis had a lower mean age (47 years) and correspondingly reduced likelihood of harboring undiagnosed neurodegenerative disease, providing higher confidence in their disease-free status.

Several limitations should be acknowledged. The current studies were performed with modest cohort sizes, and more extensive studies with larger well-characterized sample sets and comparisons against additional platforms are needed to fully establish SPLASH’s performance across diverse patient populations. Additionally, while we demonstrated proof-of-concept using DPS samples, further optimization of sample collection and extraction protocols is required before DPS-based testing can be implemented in real-world settings. Future studies should also include longitudinal validation to assess SPLASH’s utility for monitoring disease progression and treatment response.

In summary, while blood-based testing for neurodegenerative diseases has made significant strides, broad implementation remains limited by the need for specialized instrumentation and technical expertise. SPLASH addresses this barrier by leveraging standard benchtop qPCR equipment, providing potential compatibility with the large and growing global installed base of qPCR instruments across diverse research and clinical settings. With demonstrated performance across multiple sample types – including CSF, plasma, and DPS – and the ability to multiplex biomarkers with sub-pg/mL sensitivity, SPLASH is uniquely positioned to support global implementation of ultrasensitive biomarker quantification for neurodegenerative diseases.

## Materials and Methods

### Antibody Probe Synthesis and Purification

Antibody–oligonucleotide conjugates were generated using azido-based linking chemistry. Capture and detector antibodies for each target were conjugated to single-stranded oligonucleotides. Capture antibodies were additionally biotinylated using amine-reactive NHS-activated biotin. All conjugates were assessed and purified using HPLC to ensure uniformity and consistency.

### SPLASH Assay Workflow

Plasma samples were first diluted 1:4 and reference standard materials were serially diluted 1:4 using SPLASH Sample Buffer to create the standard curve. The materials were then loaded into individual wells of a 96-well qPCR plate. The samples and standard curve were then incubated with antibody probes for 2 hours at room temperature. Streptavidin-coated magnetic beads were then incubated with the mixture for 10 mins to capture the formed immunocomplexes. Washing was performed by placing the plate on a magnetic plate and removing the supernatant; 200 µL of SPLASH Wash Buffer was added and removed twice in each reaction well. A custom qPCR master mix containing ligase and qPCR probes, was then added to the magnetic beads with captured immunocomplex and placed directly in a benchtop qPCR instrument (QuantStudio 3, Thermo Fisher Scientific, Waltham, MA, USA) for ligation and signal amplification. qPCR instrument protocol is as follows:

**Table 3.** Thermocycling settings for SPLASH assay on qPCR system.

| Step | Temperature | Time | Stage |
| --- | --- | --- | --- |
| Ligation | 25°C | 10 min | Hold |
| Ligase Inactivation | 95°C | 3 min | Hold |
| PCR | 95°C | 10 s | 40 Cycles |
|  | 60°C | 30 s |  |

### Data Analysis and Concentration Calculation

Cq (quantification cycle) values from qPCR were generated using the instrument manufacturer’s analysis software. Standard curves were generated using seven concentration points and fitted using four-parameter logistic (4PL) regression. All standard curves achieved R^2^ values greater than 0.99. The resulting standard curves were used to calculate biomarker concentrations in unknown samples taking into account the 4x dilution and to convert Cq values to concentrations reported in pg/mL.

### Analytical Performance Evaluation

Six runs were performed for each assay for the estimation of the LLOD and LLOQ. For LLOD, the concentration value of the blanks (n = 15) was first calculated using the standard curve. Then the LLOD for a given run was calculated by multiplying the standard deviation of the blanks by 2.5 and adding the result to the mean of the blanks. We then took the median of the LLODs across the six runs as the overall LLOD. fLLOD was calculated by multiplying the LLOD by the dilution factor (4X).

The LLOQ is estimated by the lowest point on the standard curve for each run in which the CV was below 20% and the recovery was between 80 and 120%. The values were determined for each of the six runs. We then took the median of the LLOQs across the six runs as the overall LLOQ. fLLOQ was calculated by multiplying the LLOQ by the dilution factor (4X).

Intra-plate variability was assessed by running 2 plasma samples run as 3 or 4 replicates for each target on a single plate along with the standard curve. CV is calculated by dividing the standard deviation of the measurements by the mean of the measurements. Overall CV is the average of the CVs of the five assays.

### SPLASH-Simoa Comparison

For the platform comparison, 20 commercially sourced plasma samples 11 AD− samples (mean age 47 years) and 9 AD+ samples (mean age 75.5 years) from BioIVT (Woodbury, NY, USA) were analyzed using both SPLASH and Simoa platforms.

Plasma pTau-217 concentrations were measured on the Simoa HD-X Analyzer using the ALZpath pTau-217 Advantage V2 kit according to manufacturer’s instructions. Samples were run in triplicate for both assays.

Correlation between SPLASH and Simoa measurements was evaluated using linear regression with goodness of fit reported by R^2^. Statistical significance for group differences was determined using ANOVA followed by Kruskal-Wallis test for multiple comparisons.

### Multiplexed Biomarker Panel Evaluation

Samples for multiplexed biomarker analysis consisted of 50 commercially sourced, clinically uncharacterized samples from individuals aged 50-80 years old and 19 samples from patients with confirmed Alzheimer’s disease (BioIVT, Woodbury, NY, USA). All samples were thawed and aliquoted upon receipt and stored at -80 °C until analysis. Each plasma sample was quantified using the standard SPLASH assay workflow. The appropriate number of optical channels was selected for the triplex (three channels) and duplex (two channels) assays on the qPCR instrument (QuantStudio 3, Thermo Fisher Scientific, Waltham, MA, USA).

Biomarker classification thresholds were established using an AD-anchored approach in the absence of clinically confirmed disease-free samples.

The cutoff values were determined to yield three levels -

1. Normal Range (Blue): Values below 10th percentile of the AD-positive samples.
2. Intermediate Range (Yellow): Values that include AD-positive samples but are below the median of the AD samples.
3. Abnormal Range (Orange): Values at or exceeding/below the median of confirmed AD-positive samples.

The following classification thresholds were established:

- Aβ1–42/Aβ1–40 ratio (lower values indicate pathology): Normal: >0.37, Intermediate: 0.28-0.37, Abnormal: <0.28
- GFAP (pg/mL) (higher values indicate pathology): Normal: <17, Intermediate: 17-39, Abnormal: >39
- pTau-217 (pg/mL) (higher values indicate pathology): Normal: <0.31, Intermediate: 0.30-0.70, Abnormal: >0.70
- NfL (pg/mL) (higher values indicate pathology): Normal: <0.42, Intermediate: 0.42-1.19, Abnormal: >1.19

**Table 4.**
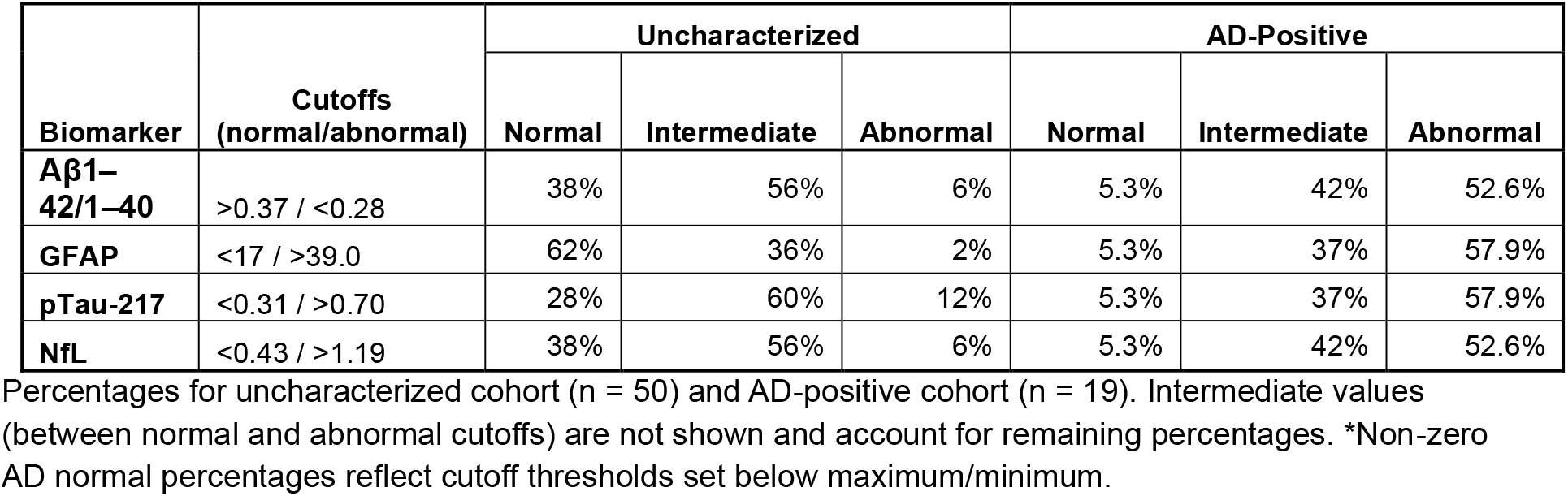
Biomarker classification of uncharacterized and AD-positive cohorts based on established cutoff values.

### DPS Sample Collection

DPS samples were collected using SEP10 cards (Capitainer, East Providence, RI, USA) from research participants recruited by the Parent Program Grant (PPG), Brain Aging Network for Cognitive Health (BrANCH), and Alzheimer’s Disease Research Center (ADRC) at the University of California, San Francisco (UCSF) Edward and Pearl Fein Memory & Aging Center (Fein MAC). All research participants or their surrogates provided written informed consent, and all research protocols were approved by the University of California, San Francisco (UCSF) institutional review board. A total of 16 participants were collected over the course of one month. DPS samples were stored at RT for 1-34 days prior to elution (mean = 19; SD = 10)

### Cohort Characteristics

For the DPS studies, the cohort comprised of clinically normal (CN, n = 6), early-onset Alzheimer’s disease (EOAD, n = 2), mild cognitive impairment (MCI, n = 2), behavioral variant frontotemporal dementia (bvFTD, n = 4), and primary progressive aphasia (PPA, n = 2) participants. There were no significant differences in age, sex, or APOE e4 status across the cohort (Table 5).

**Table 5.** Cohort demographics by diagnostic grouping.

| Variable | CN<br>(N = 6) | EOAD<br>(N = 2) | MCI<br>(N = 2) | bvFTD<br>(N = 4) | PPA<br>(N = 2) | Total<br>(N = 16) | P-value |
| --- | --- | --- | --- | --- | --- | --- | --- |
| **APOE e4 count** |  |  |  |  |  |  | 0.695 |
| 0 | 4 (66.7%) | 1 (50.0%) | 2 (100.0%) | 3 (75.0%) | 2 (100.0%) | 12 (75.0%) |  |
| 1 | 2 (33.3%) | 1 (50.0%) | 0 (0.0%) | 1 (25.0%) | 0 (0.0%) | 4 (25.0%) |  |
| 2 | 0 (0.0%) | 0 (0.0%) | 0 (0.0%) | 0 (0.0%) | 0 (0.0%) | 0 (0.0%) |  |
| **Sex** |  |  |  |  |  |  | 0.136 |
| 1 | 3 (50.0%) | 0 (0.0%) | 0 (0.0%) | 3 (75.0%) | 2 (100.0%) | 8 (50.0%) |  |
| 2 | 3 (50.0%) | 2 (100.0%) | 2 (100.0%) | 1 (25.0%) | 0 (0.0%) | 8 (50.0%) |  |
| **Age** |  |  |  |  |  |  | 0.811 |
| Mean (SD) | 67.1 (20.0) | 57.4 (2.5) | 73.0 (18.0) | 65.2 (6.4) | 58.7 (1.1) | 65.1 (13.7) |  |
| Range | 34.5 - 88.8 | 55.7 - 59.2 | 60.3 - 85.7 | 56.2 - 71.0 | 57.9 - 59.4 | 34.5 - 88.8 |  |

P-values computed with a chi-square test for categorical variables sex and APOE e4 copy, and ANOVA for continuous variable age.

### DPS Quantitation

Whole blood drawn in 8.5 mL acid-citrate dextrose (ACD) phlebotomy tubes was aliquoted (70 µL) into each two lanes of a Capitainer SEP10 card per participant, according to manufacturer’s protocol. Upon successful sampling, a consistent 10 µL volume of plasma is collected per SEP10 disc, which is then dried and stored at room temperature as a DPS until downstream processing for AD biomarker detection of pTau217 and Aβ1–42. Briefly, each DPS was eluted individually in 200 µL of SPLASH Sample Buffer (20-fold dilution) for 45 minutes under agitation. Eluates were briefly spun down, transferred to fresh tubes and held on ice until assay plating according to SPLASH standard procedure. Each SEP10 disc per sample was run in duplicate. After converting sample Cq values to concentrations (pg/ml) using a standard curve, concentrations were adjusted to account for dilution of 1.5 mL ACD per phlebotomy tube total volume, and 20-fold elution. Corrected biomarker values were used to calculate a pTau217:Aβ1–42 ratio for cohort assessment.

Coefficients of variation were calculated as the standard deviation divided by the mean, expressed as a percentage. For the pTau-217/Aβ1–42 ratio, the independent error prediction was calculated as √[CV(pTau-217)^2^ + CV(Aβ1–42)^2^].

All methods were carried out in accordance with relevant guidelines and regulations.

## Acknowledgements

J.S.Y. receives funding from NIH-NIA R01AG057234, P30AG062422, P01AG019724, and U19AG079774; NIH-NINDS U54NS123985; the Rainwater Charitable Foundation; the Alzheimer’s Association; the Global Brain Health Institute; and the Mary Oakley Foundation. The content of this publication is solely the responsibility of the authors and does not necessarily represent the official views of the NIH.

## Data Availability Statement

The datasets generated and analyzed during this study are available from the corresponding author upon reasonable request. Aggregate data supporting the findings of this study are presented in the article.

